# Identification of eight novel variants across *PAX3, SOX10, EDNRB* and *MITF* genes in Waardenburg syndrome with next-generation sequencing

**DOI:** 10.1101/2022.01.18.21267034

**Authors:** Chen-Yu Lee, Ming-Yu Lo, You-Mei Chen, Pei-Hsuan Lin, Chuan-Jen Hsu, Pei-Lung Chen, Chen-Chi Wu, Jacob Shu-jui Hsu

## Abstract

Waardenburg syndrome (WS) is a hereditary, genetically heterogeneous disorder characterized by variable presentations of sensorineural hearing impairment and pigmentation anomalies. This study aimed to investigate the clinical features of WS in detail and determine the genetic causes of patients with clinically suspected WS. A total of 24 patients from 21 Han Taiwanese families were enrolled and underwent comprehensive physical and audiological examination. We applied targeted next-generation sequencing (NGS) to investigate the potential causative variants in these patients and further validated the candidate variants through Sanger sequencing. We identified 18 causative variants of WS in our cohort. Of these variants, eight were novel and discovered in *PAX3, SOX10, EDNRB, MITF* genes, including missense, nonsense, deletion, and splice site variants. Several patients presented skeletal deformities, hypotonia, megacolon, and neurological disorders that were rarely seen in WS. This study revealed highly phenotypic variability in Taiwanese WS patients and demonstrated that targeted NGS allowed us to clarify the genetic diagnosis and extend the genetic variant spectrum of WS.

## Introduction

Waardenburg syndrome (WS) is a hereditary auditory-pigmentary disorder characterized by sensorineural hearing impairment (SNHI) and pigmentation abnormalities of the eye, hair, and skin [1]. The disease has an estimated prevalence rate of 1/42,000 [2] and accounts for 2–5% of patients with congenital hearing impairment [3].

WS is classified into four subtypes according to the presented clinical phenotype. Type 1 WS (WS1, OMIM# 193500) is diagnosed according to the criteria proposed by the Waardenburg Consortium [4] that states that a patient must have two major criteria or one major plus two minor criteria to be considered as affected. Of all the symptoms of WS, dystopia canthorum is the most distinctive feature of WS1 [2]. Type 2 WS (WS2) is distinguished from WS1 by the absence of dystopia canthorum [5]. Based on its genetic heterogeneity, WS2 is further divided into WS2A (OMIM# 193510), WS2B (OMIM% 600193), WS2C (OMIM% 606662), WS2D (OMIM# 608890), and WS2E (OMIM# 611584). Type 3 WS (WS3, OMIM# 148820), also called Klein– Waardenburg syndrome, presents as musculoskeletal abnormalities with the clinical features of WS1[6]. Type 4 WS (WS4), named Shah-Waardenburg syndrome, has similar traits to WS2 but is accompanied by Hirschsprung disease (HD, OMIM# 142623) [7]. WS4 is additionally classified into three subgroups, WS4A (OMIM# 277580), WS4B (OMIM# 613265), and WS4C (OMIM# 613266), according to genetic characteristics.

WS presents with a high degree of genetic heterogeneity. Six genes have been confirmed to cause WS: paired box 3 (*PAX3*) [8], melanocyte inducing transcription factor (*MITF*) [9, 10], SRY-box transcription factor 10 (*SOX10*) [5, 11], endothelin 3 (*EDN3*) [12], endothelin receptor type B (*EDNRB*) [13], and snail family transcriptional repressor 2 (*SNAI2*) [14]. Variants in *PAX3* are primarily responsible for WS1 and WS3, while *SOX10, EDN3*, and *EDNRB* variants are involved in WS4. Genetic studies have linked WS2 to variants in *MITF, SOX10, EDN3, EDNRB*, and *SNAI2* [15]. Interactions between the six WS-associated genes are believed to form a *MITF*-centered regulatory framework responsible for controlling the differentiation and development of neural crest cells (NCCs), particularly melanocytes derived from NCCs [15, 16]. An abnormality in this framework could lead to the development of WS and related diseases [16].

WS patients display phenotypic variability and reduced penetrance, making it difficult to diagnose the disease solely on the basis of clinical symptoms [17]. Additionally, although more than 400 pathogenic variants have been identified to cause WS [18], there are still a number of cases that are unexplained at the molecular level [17]. As such, the identification of novel variants will contribute to a better understanding of WS pathogenesis. Meanwhile, phenotypic analysis of WS patients is also crucial for better determining the correlations between genotypes and phenotypes for each WS subtype, in turn providing guidance for genetic counseling, diagnosis, and treatment choices for patients [15]. In this study, we examined in detail the clinical phenotypes in a cohort of individuals with suspected WS and performed comprehensive genetic analyses using a targeted next-generation sequencing (NGS) approach.

## Material and Methods

### Study Patients

This study included 24 patients with clinically suspected WS from 21 unrelated Han Taiwanese families. The diagnoses were made by experienced otolaryngologists or pediatricians based on the presentation of at least one major diagnostic criterium of the syndrome [4]: SNHI, blue iris, white forelock, or dystopia canthorum. Fourteen individuals with suspected WS were patients at National Taiwan University Hospital, whereas ten had been patients at ten other hospitals and were referred to National Taiwan University Hospital for genetic testing. The study was approved by the Research Ethics Committees of the National Taiwan University Hospital. All participants and/or their parents provided written informed consent.

### Characterization of Phenotypes

We obtained and analyzed the family history, medical records, and results of physical, dermatological, musculoskeletal, neurologic, and audiological examinations of all participants. Additionally, the patients underwent otoscopic and audiometric evaluations performed by experienced audiologists using pure tone audiometry or diagnostic auditory brainstem response, depending on the patient’s age and neurologic status [19, 20].

### Next-Generation Sequencing

We collected peripheral blood from the patients to extract genomic DNA from mononuclear cells. A sonication method (Covaris, Woburn, MA) was used to produce DNA fragments of an average size of 800 bp, and the length and concentration of the fragments were measured using a 2100 Bioanalyzer (Agilent Technologies, Santa Clara, CA) and Qubit (Thermo Scientific, Waltham, MA), respectively. A DNA library was constructed from the fragments using a TruSeq Library Preparation Kit (Illumina Inc., San Diego, CA). Probe capture-based target enrichment was achieved with a SeqCap EZ Hybridization and Wash Kit (Roche NimbleGen, Madison, WI) using probes designed to target the coding regions of 30 common deafness-associated genes in the Taiwanese population, including the six genes (*PAX3, MITF, EDNRB, EDN3, SOX10, SNAI2*) reported to be causative of WS (Supplemental Table S1) [15, 21]. The total target region size was approximately 317 kb, and the DNA samples were sequenced with a MiSeq platform (Illumina Inc., San Diego, CA) to produce 300-nucleotide paired-end reads with a 150x average read depth.

### Data Analyses

The paired-end reads from the NGS sequencing were aligned, sorted, and converted using the BWA-MEM and Sort utility in the Sentieon DNAseq pipeline version 2018 (https://www.sentieon.com/products/) [22]. The Sentieon Haplotyper algorithm was used to detect variants, including single nucleotide substitutions and small insertions and deletions. We used ANNOVAR version 2019 (https://annovar.openbioinformatics.org/en/latest/#annovar-documentation) [23] to annotate all of the called variants with a series of information, including Human Genome Variation Society nomenclatures, maximum allele frequency across distinguished populations of the gnomAD database [24], minor allele frequency in the Taiwan Biobank database [25], and various *in silico* prediction outcomes, including PolyPhen-2 version 2 (Harvard University, Cambridge, MA), SIFT version 2019 (SANS Institute, North Bethesda, MD), LRT version 2009 (Washington University, St. Louis, MO), MutationTaster version 2 (Charité e Universitätsmedizin Berlin, Berlin, Germany), VariantAssessor version 3 (Computational Biology Center, Memorial Sloan-Kettering Cancer Center, NY), FATHMM version 2.3 (University of Bristol, Bristol, England), and MetaLR version 2015 (Human Genetics Center, University of Texas Health Science Center at Houston, Houston, TX). We excluded variants with an allele frequency of more than 1% in both the gnomAD and Taiwan Biobank database and chose the filtered variants that were within exome or intron splicing sites. Human disease databases, including ClinVar [26] and Deafness Variation Database [27], were used to identify previously reported pathogenic variants. The online platform VarSome [28] was adopted to carefully assess all of the filtered variants in accordance with the American College of Medical Genetics and Genomics (ACMG) guidelines [29]. The variants that met the criteria for pathogenic and likely pathogenic were recorded as disease-causing and confirmed with Sanger sequencing.

## Results

### Clinical Phenotypes

A total of 24 patients from 21 unrelated families were recruited for this study (14 female and ten male patients). Four probands of the participants were clinically suspected of having WS1 (4/21, 19.0%) and seven of having WS2 (7/21, 33.3%); the other ten probands had at least one major symptom of WS but failed to fulfill the criteria for a definite clinical diagnosis [4, 7]. Genetic diagnoses were confirmed in 18 WS patients from 15 families (see “Variants Findings” below). Considering the penetrance of the causative variatns, SNHI (12/18, 66.7%) and blue iris (13/18, 72.2%) were the most frequent phenotypes among these patients. White forelock was observed in two patients (2/18, 11.1%), and dystopia canthorum was observed in four patients (4/18, 22.2%). Three patients were members of the WSF-13 family, and two other patients belonged to the WSF-14 family (Figure 2). WSF-1 presented with synophrys, broad high nasal root, and premature graying of hair, which was also observed in WSF-11. Nasolacrimal duct aplasia was discovered in WSF-2. We noted that WSF-6 exhibited hypotonia and inner ear deformity, confirmed by magnetic resonance imaging, and WSF-10 presented with megacolon. In addition, WSF-12 was reported to have facial dysmorphism and bilateral hydrocephalus. **Table 1** lists the clinical features of the 18 patients with confirmed genetic diagnoses.

**Table 1.** Summary of Phenotypic Features of 18 WS Patients

| Subjects | Gender | SNHI | Blue iris | White forelock | Dystopia canthorum | Family history* | Other abnormality | WS types |
| --- | --- | --- | --- | --- | --- | --- | --- | --- |
| WSF-1 | F | - | + | + | + | - | Synophrys, Broad high nasal root, PGH | WS1 |
| WSF-2 | F | - | + | - | + | +/-<sup>†</sup> | Nasolacrimal duct aplasia | WS1 |
| WSF-3 | F | + | - | - | - | - | - | N/A |
| WSF-4 | F | + | - | - | - | - | - | N/A |
| WSF-5 | M | - | + | - | + | - | - | WS1 |
| WSF-6 | F | + | + | - | - | - | Hypotonia, Inner ear deformity<sup>‡</sup> | WS2 |
| WSF-7 | M | + | + | - | - | - | - | WS2 |
| WSF-8 | M | + | + | - | - | - | - | WS2 |
| WSF-9-II:1 | M | + | + | - | - | - | - | WS2 |
| WSF-10 | F | - | + | - | - | - | Megacolon, Brown hair | N/A |
| WSF-11 | F | - | + | - | + | - | Constipation<sup>§</sup>, PGH | WS1 |
| WSF-12 | F | + | - | + | - | - | Facial dysmorphism, Bilateral hydrocephalus | WS2 |
| WSF-13-I:1 | M | + | + | - | - | + | - | WS2 |
| WSF-13-II:1 | F | + | + | - | - | + | - | WS2 |
| WSF-13-II:2 | M | + | - | - | - | + | - | WS2 |
| WSF-14-II:1 | M | - | + | - | - | + | - | WS2 |
| WSF-14-II:3 | F | + | + | - | - | + | - | WS2 |
| WSF-15 | M | + | - | - | - | - | - | N/A |

WS: Waardenburg syndrome; F: female; M: male; N/A: not applicable to the diagnostic criteria for WS; SNHI, sensorineural hearing impairment; PGH, premature graying of hair
\* First-degree relative previously diagnosed with WS
<sup>†</sup>WSF-2's mother had profound hearing loss but had not received genetic testing for WS.
<sup>‡</sup>WSF-6's magnetic resonant imaging showed bilateral cochlear deformity with reduced size and only 2 turns.
<sup>§</sup>WSF-11's abdominal x-ray revealed no dilated bowel loop, which made it less likely to diagnose megacolon.

### Variant Findings

Causative gene variants of *EDNRB, MITF, PAX3*, or *SOX10* were identified in eighteen patients, with 13 of the variants being unique. Eight of the variants were novel variants and presented in ten patients, and five previously reported variants were identified in eight patients (Table 2 and Supplemental Table S2) [9, 30-37]. The novel variants included missense, nonsense, and splice site changes, as well as two frameshift deletions.

**Table 2.** Causative Variants Identified in the 18 WS Patients*

| Subjects | Variant profile |  |  |  | ACMG criteria |
| --- | --- | --- | --- | --- | --- |
|  | Variants | Structure | Protein change | Zygote<sup>†</sup> |  |
| WSF-1 | *PAX3* (NM_181459.4):<br>c.807C>A, Missense | Exon | p.Asn269Lys | 1/0 | PS1, PM1, PM2, PP2, PP3 (P) |
| WSF-2 | *PAX3* (NM_181459.4):<br>c.52C>T, Nonsense | Exon | p.Gln18Ter | 1/0 | PVS1, PM2, PP3 (P) |
| WSF-3 | *PAX3* (NM_181459.4):<br>c.192C>A, Missense | Exon | p.His64Gln | 1/0 | PM1, PM2, PP2, PP3 (LP) |
| WSF-4 | *PAX3* (NM_013942.5):<br>c.587-2A>G Splice site<br>*PAX3* (NM_181459.4):<br>c.586+408A>G, Non-coding | Intron | N/A | 1/0 | PVS1, PM2 (LP, NM_013942.5)<br>PM2, BP4 (US, NM_181459.4) |
| WSF-5 | *PAX3* (NM_181459.4):<br>c.812G>A, Missense | Exon | p.Arg271His | 1/0 | PP5, PM1, PM2, PM5, PP2, PP3 (P) |
| WSF-6 | *SOX10* (NM_006941.4):<br>c.684_693del, Deletion | Exon | p.Glu229Glnfs<br>Ter54 | 1/0 | PVS1, PM2, PP3 (P) |
| WSF-7 | *SOX10* (NM_006941.4):<br>c.314_315del, Deletion | Exon | p.Lys105Thrfs<br>Ter28 | 1/0 | PVS1, PM2, PP3 (P) |
| WSF-8 | *SOX10* (NM_006941.4):<br>c.424T>G, Missense | Exon | p.Trp142Gly | 1/0 | PM1, PM2, PM5, PP2, PP3 (P) |
| WSF-9-II:1 | *EDNRB* (NM_001201397.1):<br>c.754-2A>G, Splice site | Intron | N/A | 1/0 | PVS1, PM2, PP3 (P) |
| WSF-10 | *EDNRB* (NM_001201397.1):<br>c.823G>A, Missense | Exon | p.Val275Met | 1/1 | PM1, PM2, PP3 (US) |
| WSF-11 | *EDNRB* (NM_001201397.1):<br>c.823G>A, Missense | Exon | p.Val275Met | 1/1 | PM1, PM2, PP3 (US) |
| WSF-12 | *EDNRB* (NM_001201397.1):<br>c.823G>A, Missense | Exon | p.Val275Met | 1/0 | PM1, PM2, PP3, (US) |
| WSF-13-I:1 | *MITF* (NM_198159.3):<br>c.1052C>T, Missense | Exon | p.Ser351Phe | 1/0 | PM1, PM2, PM5, PP2, PP3 (P) |
| WSF-13-II:1 | *MITF* (NM_198159.3):<br>c.1052C>T, Missense | Exon | p.Ser351Phe | 1/0 | PM1, PM2, PM5, PP2, PP3 (P) |
| WSF-13-II:2 | *MITF* (NM_198159.3):<br>c.1052C>T, Missense | Exon | p.Ser351Phe | 1/0 | PM1, PM2, PM5, PP2, PP3 (P) |
| WSF-14-II:1 | *MITF* (NM_198159.3):<br>c.1078C>T, Nonsense | Exon | p.Arg360Ter | 1/0 | PVS1, PM2, PP3, PP5 (P) |
| WSF-14-II:3 | *MITF* (NM_198159.3):<br>c.1078C>T, Nonsense | Exon | p.Arg360Ter | 1/0 | PVS1, PM2, PP3, PP5 (P) |
| WSF-15 | *MITF* (NM_198159.3):<br>c.1066C>T, Nonsense | Exon | p.Arg356Ter | 1/0 | PVS1, PM2, PP3, PP5 (P) |

\*For detailed information about the reporting history and allele frequency of the identified variants, please refer to the **Supplemental Table S2**.
<sup>†</sup>1/0: heterozygous, 1/1: alternative homozygous
ACMG, American College of Medical Genetics and Genomics; N/A, not applicable; P, pathogenic; LP, likely pathogenic; US, uncertain significance

Of the eight novel variants, two missense variants and one nonsense variant of *PAX3* were detected: NM_181459.4:c.192C>A in the paired domain, c.807C>A in the homeodomain, and c.52C>T in exon 1. Three heterozygous variants in *SOX10* were identified: NM_006941.4:c.314_315del and c.424T>G in the high mobility group (HMG) domain and a frameshift deletion c.684_693del between the HMG and K2 domains. Additionally, the *EDNRB* variant NM_001201397.1:c.754-2A>G occurred in the splice site located 2 bp upstream of exon 3, and a novel *MITF* heterozygous missense variant NM_198159.3:c.1052C>T was identified, which could cause an amino acid replacement in the homeodomain of MITF.

All ten reported variants were either included in ClinVar [26] or Deafness Variation Database [27], or both. The *PAX3* heterozygous variant c.812G>A had been documented in 21 studies according to VarSome [28]. Research about the *EDNRB* homozygous variant c.823G>A was reported in Taiwan [30], China [32], and Pakistan [31]. Moreover, in three Chinese studies [9, 34, 35] and one Brazilian study [33], the *MITF* nonsense variant c.1066C>T was described. We confirmed each causative variant through Sanger sequencing. **Table 2** and **Figure 1** present the disease-causing variants identified in this study.

**Figure 1.**
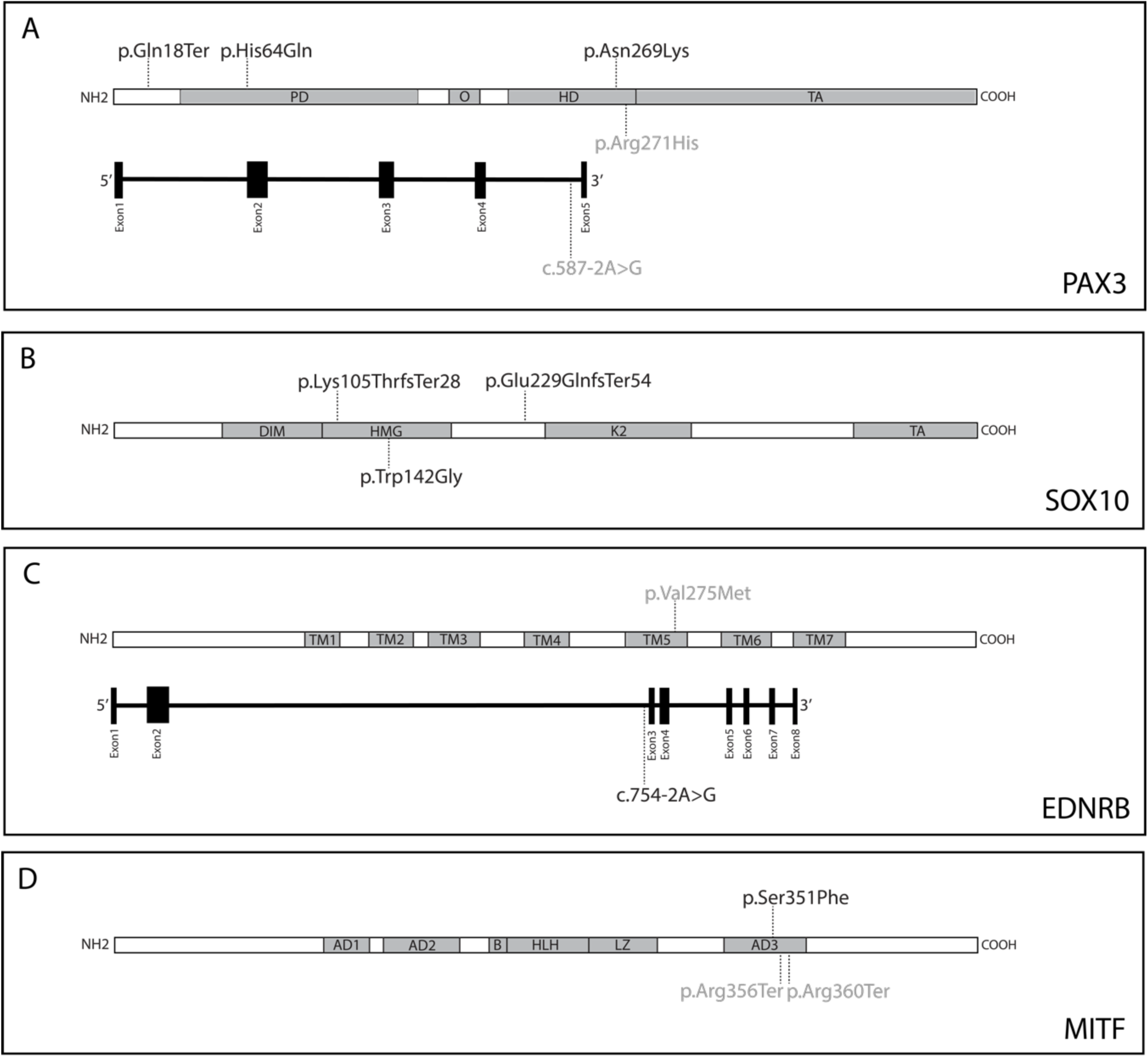
Localization of 13 causative variants in 18 diagnosed WS patients. Novel variants were shown in black and reported variants were shown in gray. A) *PAX3*: P, paired domain; O, octapeptide; HD, homeodomain; TA, transactivation domain B) *SOX10*: DIM, dimerization domain; HMG, high mobility group; K2, context-dependent transactivation domain; TA, transactivation domain C) *EDNRB*: TM, transmembrane domain D) *MITF*: AD, transactivation domain; B, basic domain; HLH, helix-loop-helix domain; LZ, leucine zipper domain

**Figure 2.**
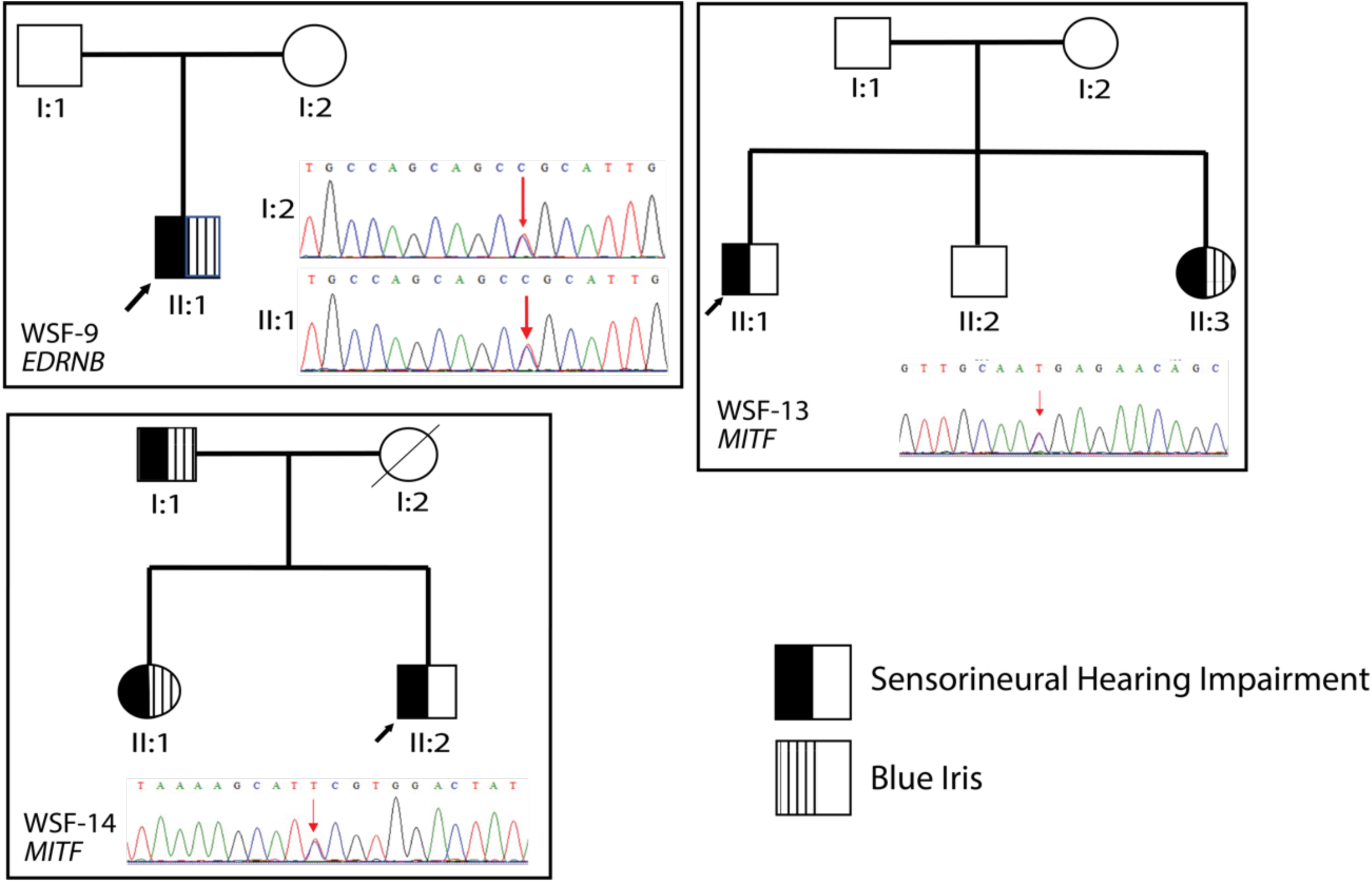
Pedigree of three families with Waardenburg syndrome. The probands were indicated by arrows. All the affected individuals in the same family shared the identical variant confirmed by Sanger sequencing. WSF-9-I:2 was not affected while having the same variant as WSF-9-II:1

## Discussion

In this study, we identified 18 variants responsible for WS in 18 clinically suspected WS patients through targeted NGS (Table 2). Of these variants, 15 were pathogenic or likely pathogenic and located in the *PAX3, SOX10, EDNRB*, or *MITF* genes. Eight variants were novel. While the same missense variant of unknown significance (*EDNRB*:c.823G>A) was harbored by three unrelated patients (WSF-10, WSF-11, and WSF-12), we concluded that the variant might be disease-causing based on molecular analysis and previous case reports (discussed below).

*PAX3* encodes a paired box transcription factor (TF) involved in the development of melanocytes through the regulation of MITF and other TFs [38, 39]. Mice with heterozygous *PAX3* variants might present with patchy loss of pigmentation, while those with homozygous variants die during pregnancy or shortly after birth [17]. In humans, *PAX3* variants are the most prevalent cause of WS [15, 17] and are responsible for most WS1 and WS3 cases [8, 15]. Homozygous or compound heterozygous *PAX3* variants might induce more severe manifestations: extended depigmentation, upper limb defects, and even death in early infancy or *in utero*, for which WS3 is likely to be diagnosed [40, 41]. In addition, the occurrence of *de novo* variants and germline mosaicism were found in some WS1 cases [8]. No apparent correlation between the variant type, location, or severity of the phenotype was observed in WS associated with *PAX3* variants, suggesting that the pathophysiology of the disease may be due to a gene dosage effect [17].

In this study, we identified three novel *PAX3* variants. *PAX*:c.52C>T was located at exon 1 and might cause truncation of the translated protein. Variant *PAX*:c.192C>A was located at the paired domain (PD), which affects DNA binding capacity as well as the transcriptional regulation of downstream target genes [17]. *PAX3*:c.807C>A was located in the homeodomain (HD) and affects the interaction of PAX3 with DNA by destabilizing the domain fold [42] (Figure 1A).

Our study also revealed that the five patients with *PAX3* variants displayed diverse phenotypes. Aside from the major diagnostic criteria for WS, WSF-1 exhibited synophrys, broad high nasal roots, and premature graying of the hair. A similar finding to the nasolacrimal duct aplasia noted in WSF-2 was reported by David and Warin [43], wherein congenital blockage of the nasolacrimal duct was observed in an 11-year old WS boy. More investigation is required to determine whether the abnormal development of the lacrimal duct is related to the dysmorphic effect that *PAX3* variants may have on the craniofacial bone [44]. In addition, both WSF-3 and WSF-4 showed isolated SNHI, while WSF-5 had only blue iris and dystopia canthorum. These findings in our patients with *PAX3* variants were consistent with the observation that the penetrance of each clinical feature of WS1 is not complete [17], indicating that genetic backgrounds and environmental factors may influence the disease manifestations [45].

*SOX10* encodes a TF critical to the early development of neural crest stem cells by promoting cell survival and maintaining multipotency [46, 47]. Moreover, SOX10 controls the development of melanocytes by regulating *MITF* expression in synergy with PAX3 [11]. Through modulation of *RET* expression, SOX10 determines the morphogenesis of the enteric nervous system (ENS) [48]. Mutant *Sox10* heterozygous mice demonstrated variability of aganglionosis, the characteristic HD phenotype in WS4 patients [49]. In humans, *SOX10* variants account for ∼15% of WS2 cases and 40–50% of WS4 patients [1]. This study revealed three novel variants in *SOX10*. Variants c.314_315del and c.424T>G were located in the highly conserved HMG domain of SOX10 that may cause conformational change and result in a decreased DNA-binding capacity [50]. In addition, *SOX10*:c.684_693del was identified in exon 3 of the gene, which activates nonsense-mediated RNA decay (NMD), leading to haploinsufficiency [17] (Figure 1B).

SNHI was exhibited in all three patients with *SOX10* variants. In a review of 417 WS patients [51], SNHI was the most common feature of WS2 (100%) and WS4 (92.9%) in patients with *SOX10* variants. Absent migration of NCC-derived melanocytes to the stria vascularis due to *SOX10* variants might impair ionic concentration of endolymph, which leads to the resultant SNHI [52]. Moreover, cochlear hypoplasia and the absence of the cochlear nerve were reported in patients with *SOX10* variants [53]. However, we only found WSF-6 to have inner ear malformation of reduced cochlear size and turns. This finding demonstrates that while *SOX10* variants might directly induce inner ear deformity [50], the phenotypic expression might be variable with incomplete penetrance, highlighting the difficulty of predicting inner ear malformations only with genotypic information.

Interestingly, besides typical manifestations of WS (SNHI and blue iris), WSF-6 also showed hypotonia without HD. Some WS4 patients with *SOX10* variants were reported to have PCWH syndrome (OMIM# 609136), a complex neurocristopathy including peripheral demyelinating neuropathy, central demyelinating leukodystrophy, Waardenburg syndrome, and Hirschsprung disease [54]. Patients with severe cases exhibit hypotonia, arthrogryposis, or respiratory insufficiency [55, 56], while those with mild cases likely display variable hypotonia, spasticity, ataxia, and developmental delay [54, 57]. To date, several WS2 patients have also presented with neuromuscular features reminiscent of PCWH [1], thus delineating a new extended PCW phenotype [17] similar to that observed in WSF-6.

EDNRB binds EDN3 and transmits signals through the Gαq/α11subunits of G-protein [58]. The signaling pathway is crucial to NCC development during the embryonic stage of vertebrates, including melanoblast differentiation and the formation of neurons and glial cells in the ENS [15, 17]. Mutant *Ednr3* mice demonstrate hypopigmentation and the absence of enteric ganglia in the distal part of the gut [55, 58]. The phenotypes observed in mutant *Ednr3* mice were similar to those in WS4 patients, in whom homozygous *EDNRB* variants were first described [59]. In contrast, heterozygous *EDNRB* variants were reported in patients with isolated HD [60, 61]. However, several studies have also proposed that the homozygous variants in *EDNRB* could induce isolated HD, and heterozygous variants might occur in WS4 [12, 52]. These findings suggest that *EDNRB* variants manifest in a more complex manner than merely as dominant or recessive conditions. Moreover, we might not be able to accurately forecast whether an *EDNRB* variant is more associated with isolated HD or WS because of the contribution of modifier genes to the development of both diseases [58].

In humans, *EDNRB*/*EDN3* variants in WS2 patients make up less than 5% of cases and in WS4 patients, 20–30% of cases [17]. We identified four patients with *EDNRB* variants. WSF-9-II:1 had a novel heterozygous splice-site variant *EDNRB*:c.754-2A>G two bp upstream of exon 3 (Figure 1C), which may lead to the absence of the genetic product [29]. While WSF-9-II:1 was diagnosed with WS2, his mother, who harbored the same heterozygous variant, was not affected (Figure 2). WSF-10 and WSF-11 harbored the same homozygous *EDNRB*:c.823G>A variant (Table 2), which was initially identified in two Chinese patients and one Taiwanese individual in the heterozygous form with isolated HD [30, 32]. However, WSF-10 and WSF-11 showed distinct phenotypes of WS. WS10 had megacolon and blue iris; WSF-11 exhibited blue iris, dystopia canthorum, premature graying of hair, and constipation (Table 1). Additionally, we found that although WSF-12 had heterozygous *EDNRB*:c.823G>A, she did not present with any gastrointestinal symptoms but possessed the phenotypes of WS2, facial dysmorphism, and bilateral hydrocephalus. Few cases of WS1 and WS4 were observed to have hydrocephalus, indicating WS increases the risk of neural tube defects in WS [62] and pathologic association with other developmental disorders [63]. Clinically, WSF-10 did not meet the current diagnostic criteria of WS4 due to only presenting with blue iris and megacolon; WSF-11 was diagnosed as WS1, which is rarely caused by *EDNRB* variants [64, 65]. These findings demonstrate the highly variable nature of phenotypic expression and the reduced penetrance of *EDNRB* features.

Additionally, we identified the missense variant *EDNRB*:c.823G>A located in transmembrane domain 5 (TM5, Figure 1C) [15] with different interpretations of pathogenicity in ClinVar (conflicting interpretation), DVD (pathogenic), and VarSome (uncertain significance) (Table 2). The missense variants in TM5 of EDNRB were reported to cause dysfunctional intracellular signaling and impair the translocation of receptors to the plasma membrane [66, 67]. Together with the previous case reports [30-32], these findings could lead to the interpretation of this variant being modified from an undetermined significance to likely disease-causing.

MITF is a basic helix-loop-helix zipper (bHLH-Zip) TF that belongs to the Myc supergene family and regulates gene expression by functioning as a homo-or heterodimeric TF [68]. MITF is extensively involved in the regulation and signaling pathways associated with the development of melanocytes and retinal pigment epithelial cells (RPE) [16]. *Mitf* mutant mice display depigmented coat colors (reduced, spotty, or absent), and some of them might present with deafness and small or absent eyes because of impaired RPE morphogenesis [16, 68]. The absence of intermediate cells and a decrease in cochlear potential in the stria vascularis were observed in mutant *Mitf* mice, thus possibly explaining the SNHI phenotype in patients with *MITF* variants [15]. In humans, about 15% of WS2 patients possess *MITF* variants [2]. Some studies have described variable phenotypes and incomplete penetrance of each symptom in multiplex families [10, 69], as was observed in the WSF-13 and WSF-14 families (Figure 2). The mild or absent manifestations of WS in individuals with *MITF* variants suggest that other modifying factors may affect the disease’s expression [10]. In our study, the novel missense *MITF* variant c.1052C>T caused an amino acid change in the third transactivation domain (AD3, Figure 1D) and was responsible for WS in the WSF-13 family.

In this study, we demonstrated the strength of NGS for identifying novel variants in WS-associated genes. Moreover, we also revealed that WSF-19 (*GJB2*), WSF-20 (*SLC26A4*), and WSF-21 (*OTOF*) harbored variants associated with SNHI-related genes (Supplemental Table S3), validating the use of NGS to clarify the underlying mechanism of isolated SNHI [70]. However, this study had several limitations. First, we noted that three patients could not be genetically diagnosed with WS or other diseases. No candidate variant was detected in WSF-16, indicating that the causative variant may reside in regions other than the six WS-associated genes [71], the presence of structural variants [33, 72], or the existence of variants in non-coding regions [73], detection of which were beyond the limits of the NGS technique used in this study. Variants of uncertain significance were recognized in WSF-17 (*PAX3*) and WSF-18 (*MITF*), suggesting that further functional study may be required to elucidate the exact effect of these variants. Second, we recruited a relatively small cohort size of 24 patients from a single population (Han Taiwanese), limiting the generalizability of the study results. **Supplemental Table S3** lists the phenotype and genotype information of six patients unable to be genetically diagnosed.

In conclusion, our results support that NGS is a useful procedure for diagnosing WS. With the help of NGS, we identified eight novel variants in *SOX10, EDNRB, MITF*, and *PAX3* that may cause WS in Taiwanese patients. Furthermore, we validated the characteristics of incomplete penetrance and variable phenotypes in WS, suggesting that a more complex regulatory framework may govern the genetic expression of the disease.

## Data Availability

All data produced in the present study are available upon reasonable request to the authors
All data produced in the present work are contained in the manuscript
All data produced are available online at

## Author Contributions

C.-Y.L. designed and directed the study and drafted the manuscript; Y.-M.C. performed the NGS experiment; M.-Y.L. performed the Sanger sequencing; C.-Y.L., M.-Y.L., Y.-M.C. contributed equally to the data analyses; P.-H.L., C.-J.H., P.-L.C., C.-C.W. provided clinical resources and advices; P.-L.C., C.-C.W., S.-J.H. critically reviewed the manuscript; S.-J.H. supervised the whole research.

## Ethics Approval

This study was performed in line with the principles of the Declaration of Helsinki. Approval was granted by the Research Ethics Committees of the National Taiwan University Hospital (2021/11/09, No. 1103705165). All participants and/or their parents provided written informed consent.

## Consent to Publish

The authors affirm that human research participants provided informed consent for publication of the data related to their phenotypes and genotypes.

## Acknowledgements

We would like to thank all subjects and their family members for participating in the present study. We also wish to thank National Center for High-performance Computing (NCHC) of National Applied Research Laboratories (NARLabs) in Taiwan for providing computational and storage resources.

