## Supplemental Table S1 for "Identification of eight novel variants across *PAX3, SOX10, EDNRB* and *MITF* genes in Waardenburg syndrome with next-generation sequencing"

| **Supplemental Table S1** 30 Common Deafness-Associated Genes in Taiwanese Population | | | | | | | |
| --- | --- | --- | --- | --- | --- | --- | --- |
| *AIFM1* | *DFNB59* | *DIAPH3* | *EDN3* | *EDNRB* | *EYA1* | *FOXI1* | *GJA1* |
| *GJB1* | *GJB2* | *GJB3* | *GJB4* | *GJB6* | *KCNJ10* | *KCNQ4* | *MITF* |
| *PAX3* | *PCDH9* | *MTRNR1* | *MYO15A* | *OTOF* | *POU3F4* | *POU4F3* | *SIX5* |
| *SIX1* | *SLC26A4* | *SNAI2* | *STRC* | *SOX10* | *TMPRSS3* |  |  |
