## Supplemental Table S2 for "Identification of eight novel variants across *PAX3, SOX10, EDNRB* and *MITF* genes in Waardenburg syndrome with next-generation sequencing"

| **Supplemental Table S2** Causative Variants Identified in the 18 WS Patients | | | | | | | | | | | | |
| --- | --- | --- | --- | --- | --- | --- | --- | --- | --- | --- | --- | --- |
|  | Variant profile | | | |  | Variant report | |  | MAF | |  |  |
| Subjects | Variants | Structure | Protein change | Zygote* |  | Clinvar | DVD |  | gnomAD^†^ | TB^‡^ |  | ACMG  criteria |
| WSF-1 | *PAX3* (NM_181459.4):  c.807C>A, Missense | Exon | p.Asn269Lys | 1/0 |  | N/A | N/A |  | N/A | N/A |  | PS1, PM1, PM2, PP2, PP3 (P) |
| WSF-2 | *PAX3* (NM_181459.4): c.52C>T, Nonsense | Exon | p.Gln18Ter | 1/0 |  | N/A | N/A |  | N/A | N/A |  | PVS1, PM2, PP3 (P) |
| WSF-3 | *PAX3* (NM_181459.4): c.192C>A, Missense | Exon | p.His64Gln | 1/0 |  | N/A | N/A |  | N/A | N/A |  | PM1, PM2, PP2, PP3 (LP) |
| WSF-4 | *PAX3* (NM_013942.5): c.587-2A>G Splice site  *PAX3* (NM_181459.4): c.586+408A>G, Non-coding | Intron | N/A | 1/0 |  | N/A | 2:223158478: T>C (US) |  | 0.0001087 | N/A |  | PVS1, PM2 (LP, NM_013942.5)  PM2, BP4 (US, NM_181459.4) |
| WSF-5 | *PAX3* (NM_181459.4): c.812G>A, Missense | Exon | p.Arg271His | 1/0 |  | VCV000279964.3 (P) | 2:223086087: C>T (P) |  | 0 | N/A |  | PP5, PM1, PM2, PM5, PP2,PP3 (P) |
| WSF-6 | *SOX10* (NM_006941.4): c.684_693del, Deletion | Exon | p.Glu229GlnfsTer54 | 1/0 |  | N/A | N/A |  | N/A | N/A |  | PVS1, PM2, PP3 (P) |
| WSF-7 | *SOX10* (NM_006941.4): c.314_315del, Deletion | Exon | p.Lys105ThrfsTer28 | 1/0 |  | N/A | N/A |  | N/A | N/A |  | PVS1, PM2, PP3 (P) |
| WSF-8 | *SOX10* (NM_006941.4): c.424T>G, Missense | Exon | p.Trp142Gly | 1/0 |  | N/A | N/A |  | N/A | N/A |  | PM1, PM2, PM5, PP2,PP3 (P) |
| WSF-9-II:1 | *EDNRB* (NM_001201397.1): c.754-2A>G, Splice site | Intron | N/A | 1/0 |  | N/A | N/A |  | N/A | N/A |  | PVS1, PM2, PP3 (P) |
| WSF-10 | *EDNRB* (NM_001201397.1): c.823G>A, Missense | Exon | p.Val275Met | 1/1 |  | VCV000619136.3^§^ | 13:78477673: C>T(P) |  | 0.001305 | 0.002307 |  | PM1, PM2, PP3, BP1 (US) |
| WSF-11 | *EDNRB* (NM_001201397.1): c.823G>A, Missense | Exon | p.Val275Met | 1/1 |  | VCV000619136.3^§^ | 13:78477673: C>T(P) |  | 0.001305 | 0.002307 |  | PM1, PM2, PP3, BP1 (US) |
| WSF-12 | *EDNRB* (NM_001201397.1): c.823G>A, Missense | Exon | p.Val275Met | 1/0 |  | VCV000619136.3^§^ | 13:78477673: C>T(P) |  | 0.001305 | 0.002307 |  | PM1, PM2, PP3, BP1 (US) |
| WSF-13-I:1 | *MITF* (NM_198159.3): c.1052C>T, Missense | Exon | p.Ser351Phe | 1/0 |  | N/A | N/A |  | N/A | N/A |  | PM1, PM2, PM5, PP2,PP3 (P) |
| WSF-13-II:1 | *MITF* (NM_198159.3): c.1052C>T, Missense | Exon | p.Ser351Phe | 1/0 |  | N/A | N/A |  | N/A | N/A |  | PM1, PM2, PM5, PP2,PP3 (P) |
| WSF-13-II:2 | *MITF* (NM_198159.3): c.1052C>T, Missense | Exon | p.Ser351Phe | 1/0 |  | N/A | N/A |  | N/A | N/A |  | PM1, PM2, PM5, PP2,PP3 (P) |
| WSF-14-II:1 | *MITF* (NM_198159.3): c.1078C>T, Nonsense | Exon | p.Arg360Ter | 1/0 |  | VCV000995922.1 (P) | 3:70008488:C>T (P) |  | N/A | N/A |  | PVS1, PM2, PP3, PP5 (P) |
| WSF-14-II:3 | *MITF* (NM_198159.3): c.1078C>T, Nonsense | Exon | p.Arg360Ter | 1/0 |  | VCV000995922.1 (P) | 3:70008488:C>T (P) |  | N/A | N/A |  | PVS1, PM2, PP3, PP5 (P) |
| WSF-15 | *MITF* (NM_198159.3): c.1066C>T, Nonsense | Exon | p.Arg356Ter | 1/0 |  | VCV000372755.7 (LP) | 3:70008476:C>T (P) |  | N/A | N/A |  | PVS1, PM2, PP3, PP5 (P) |

* 1/0: heterozygous, 1/1: alternative homozygous;

^†^ In this study, we adopted the minor allele frequency of East Asian population from gnomAD v2.1.1 with reference genome GRCh37

^‡^ Taiwan Biobank (last accessed November 5, 2021), is the largest whole genome sequencing database in Taiwan. The official website and browser are [*https://www.twbiobank.org.tw/new_web_en/index.php*](https://www.twbiobank.org.tw/new_web_en/index.php) and [*https://taiwanview.twbiobank.org.tw/browse38*](https://taiwanview.twbiobank.org.tw/browse38)*,* respectively. The reference genome in was GRCh37.

^§^This variant had conflicting interpretation of pathogenicity on ClinVar with one Likely Pathogenic and one Uncertain Significance.

MAF, minor allele frequency; DVD, Deafness Variation Database, Version 9; TB, Taiwan Biobank; ACMG, American College of Medical Genetics and Genomics; N/A, not applicable; P, pathogenic; LP, likely pathogenic; US, uncertain significance
