## Supplemental Table S3 for "Identification of eight novel variants across *PAX3, SOX10, EDNRB* and *MITF* genes in Waardenburg syndrome with next-generation sequencing"

| **Supplemental Table S3** Phenotype and Genotype Information of Six Genetically Undiagnosed Patients | | | | | | |
| --- | --- | --- | --- | --- | --- | --- |
| Subjects | Gender | Phenotypic features | WS types | Variants | Zygosity | ACMG criteria |
| WSF-16 | F | Microtia, Brown iris, White hair in the parietal area | N/A | N/A^*^ | N/A | N/A |
| WSF-17 | F | SNHI | N/A | *PAX3* (NM_181459.4): c.1130C>G (p.Ser377Cys) | 1/0 | PM2, PP2, PP3 (US) |
| WSF-18 | M | SNHI | N/A | *MITF* (NM_198159.3): c.682G>A (p.Asp228Asn) | 1/0 | PM1, PP2, PP3, BS2 (US) |
| WSF-19 | M | SNHI, Abnormal iris color | N/A | *GJB2* (NM_004004.6): c.109G>A (p.Val37Ile) | 1/0 | PS1, PM1, PM5, PP2, PP5(P) |
| WSF-20 | F | SNHI | N/A | *PAX3* (NM_181459.4): c.71G>T (p.Gly24Val)  *SLC26A4* (NM_000441.2): c.919-2A>G | 1/0;  1/0 | PM2, PP2, PP3, BP6 (US, *PAX3*:c.71G>T)  PVS1, PP5, PS3, PM2, PP3 (P, *SLC26A4*: c.919-2A>G) |
| WSF-21 | F | SNHI | N/A | *PAX3* (NM_181459.4): c.71G>T (p.Gly24Val)  *OTOF* (NM_194248.3): c.5098G>C (p.Glu1700Gln) | 1/0;  1/0 | PM2, PP2, PP3, BP6 (US, *PAX3*:c.71G>T)  PM2, PP3 (US, *OTOF*: c.5098G>C) |

N/A, not applicable; SNHI, sensorineural hearing impairment; ACMG, American College of Medical Genetics and Genomics; P, pathogenic; LP, likely pathogenic; US, unknown significance

^*^We did not detect any possible causative variant in this patient.
